# In-Context Learning with Large Language Models for Scalable Glycemic Index Assignment to Food Composition Databases: Development, Validation, and Reproducibility

**DOI:** 10.64898/2026.04.23.26351292

**Authors:** Karen A. Della Corte, Joshua L. Ebbert, Jennie Brand-Miller, Fiona Atkinson, Dennis Della Corte

**Affiliations:** Department of Nutrition, Dietetics, and Food Science, Brigham Young University, Provo, UT, USA; Department of Physics and Astronomy, Brigham Young University, Provo, UT, USA; School of Life and Environmental Sciences and Charles Perkins Centre, University of Sydney, Sydney, Australia

**Author notes:** **Corresponding author:** Dennis Della Corte, PhD, Department of Physics and Astronomy, Brigham Young University, Provo, UT 84602, USA.

**Keywords:** glycemic index, large language model, in-context learning, food composition database, nutritional epidemiology, reproducibility, artificial intelligence

## Abstract

Assigning glycemic index (GI) values to food composition databases is a critical bottleneck in nutritional epidemiology. We developed an in-context learning approach using large language models (LLMs), in which a structured knowledge system (termed a skill) loads GI reference databases (∼11,000 entries), expert decision rules, and error-correction heuristics into the model’s context window (∼300,000 tokens). The LLM performs GI assignments without scripted logic, functioning simultaneously as a semantic matching engine, numerical reasoning system, and expert curator.

We validated this approach in two experiments. In Validation Study 1, the skill predicted the expert-curated US National GI Database (9,428 foods) using only European reference data, achieving within ±10 agreement of 73.7% without manual review – compared with 31.3% retention of previously published cosine-similarity approach. In Validation Study 2, the skill was augmented with US GIDB and applied to 1,157 European food descriptions classified using the EFSA FoodEx2 system, achieving ICC = 0.79 with the expert (weighted κ = 0.65; triplicate ICC = 0.88). We then applied the skill prospectively to extend US dietary GI and GL surveillance to two additional NHANES cycles (2019–2023), identifying a continued decline in energy-adjusted glycemic load.

Reproducibility was assessed through triplicate runs (temperature = 0, pinned model version). The skill architecture is described in sufficient detail to inform future applications of in-context learning for nutritional database construction.

**STATEMENT OF SIGNIFICANCE:** This paper introduces a fundamentally new approach to glycemic index (GI) database construction. Rather than using programmatic text-matching algorithms followed by extensive manual curation, we demonstrate that a large language model (LLM), when loaded with the complete GI reference literature and formalized expert decision rules, can perform one-shot GI assignments at accuracy levels comparable to human expert ratings (ICC = 0.79 with expert, weighted κ = 0.65 for GI category agreement). The approach is validated across two independent food databases spanning US and European food supplies. The method reduces the time required to assign GI values to a new national food database from months of expert labor to hours of computation, while maintaining reproducibility through a structured, versionable skill architecture. This has immediate practical implications for enabling GI-based dietary surveillance and epidemiologic research in countries that currently lack GI databases or need to update existing databases.

## INTRODUCTION

The glycemic index (GI) is a physiological metric that ranks carbohydrate-containing foods by their effect on postprandial blood glucose concentrations relative to a reference food (1, 2). Since its introduction by Jenkins et al. in 1981, the GI has become a central tool in nutritional epidemiology, with large prospective studies linking habitual consumption of high-GI diets to increased risk of type 2 diabetes (3), cardiovascular disease (4), and certain cancers (5). The International Tables of Glycemic Index and Glycemic Load Values, most recently updated in 2021 by Atkinson et al. (6), now catalog over 4,000 food items tested under standardized conditions, providing the foundational reference data for GI-based research.

Despite the established importance of GI in diet-disease relationships, a persistent methodological bottleneck limits its application in large-scale epidemiologic studies: the assignment of GI values to national food composition databases. National dietary surveillance systems such as the US National Health and Nutrition Examination Survey (NHANES) record food intake using country-specific food coding systems (e.g., USDA Food and Nutrient Database for Dietary Studies [FNDDS]) that contain thousands of unique food codes. Matching these codes to GI values from the International Tables requires reconciling differences in food nomenclature, preparation methods, regional food variants, and compositional ambiguities – a process that has historically demanded months of expert labor (7–10).

In our previous work (11), we developed the first nationally representative US GI database by using an artificial intelligence (AI)-enabled text-embedding approach (cosine similarity of OpenAI embeddings) to generate initial matches between USDA food codes and GI reference values, followed by extensive manual curation guided by substantive domain expertise. While this approach successfully assigned GI values covering 99.9% of total carbohydrate intake across 10 NHANES cycles (1999–2018), the initial AI accuracy was 75.0%, with only 31.3% of automated matches retained after expert review. The remaining 68.7% required manual correction – underscoring that text-similarity algorithms, while useful for initial screening, lack the domain knowledge to handle the nuanced decisions inherent in GI assignment (e.g., recognizing that meats assigned GI = 70 in the Diogenes database are methodological placeholders, not measured values; or that cooking method and grain variety systematically alter GI).

Recent advances in large language models (LLMs) have opened new possibilities for food classification and nutritional assessment tasks. Studies have demonstrated that LLMs can classify foods into nutritional categories with 90–94% agreement with human experts (12), decompose composite ingredients for meal planning (13), and estimate macronutrient content from food descriptions and images (14, 15). Domain-specialized models such as FoodyLLM have shown that incorporating nutrition-specific training data substantially improves performance over general-purpose models (16). However, no study has applied LLMs to the specific problem of GI value assignment – a task that requires not only semantic food matching but also quantitative reasoning about carbohydrate composition, expert judgment about data quality, and knowledge of GI methodology.

A key concern in applying LLMs to scientific data tasks is reproducibility. LLMs generate text by sampling from probability distributions, introducing stochastic variability that can produce different outputs for identical inputs (17–19). While recent work has proposed frameworks for quantifying LLM repeatability and reproducibility (20) and has identified engineering solutions for achieving deterministic inference (21), the implications for structured data-assignment tasks in nutritional epidemiology have not been systematically evaluated.

In this paper, we introduce a novel *skill-based in-context learning (ICL)* approach for GI assignment. A *skill* is a structured prompt that loads the complete GI reference literature – the Sydney International Tables of GI Values and the Diogenes European food GI database (∼11,000 entries combined) – alongside formalized expert decision rules, data-quality correction heuristics, and a matching hierarchy directly into the LLM’s context window (∼300,000 tokens). The model then performs GI assignments without any scripted logic, functioning simultaneously as a semantic matching engine (understanding food descriptions across languages and naming conventions), a numerical aggregation system (synthesizing values across multiple reference entries), and an expert curator (applying domain-specific error-correction rules). Effective integration of AI into nutrition research requires interdisciplinary teams in which domain expertise informs the modeling process from inception, not post hoc (22). This work embodies that principle: sustained collaboration between GI methodology experts and computational researchers produced a skill architecture that encodes decades of domain reasoning directly into the LLM’s inference framework. We validate this approach in two independent experiments, assess reproducibility through triplicate concordance analysis, and demonstrate practical utility by extending nationally representative US dietary GI surveillance to recent NHANES cycles.

## METHODS

### Overview of the Skill-Based Approach

The GI Assignment Skill is a structured text document (∼8,000 words) that encodes three categories of information loaded entirely into the LLM’s context window at inference time:

#### (1) Reference GI data

The complete Sydney International Tables of GI Values (6, 23, 24) of ∼4,200 entries and the Diogenes European food GI database (∼6,800 entries), totaling ∼11,000 unique reference food–GI pairs. These are loaded as structured CSV data, preserving food descriptions, GI values, methodology notes, and data-quality indicators (25).

#### (2) Expert decision rules

A formalized hierarchy of assignment logic, including: (a) a zero-carbohydrate rule assigning GI = 0 to single-ingredient foods with negligible available carbohydrate (e.g., unprocessed meats, fats, plain cheese, water), while retaining measured GI values for low-carbohydrate foods with published test data (e.g., dairy, some fruits); (b) a matching hierarchy specifying preference order (direct match → close match → weighted ingredient GI → category-based default → composite expert reasoning); (c) rules for handling multiple reference entries (trimmed mean with outlier exclusion); (d) a mixed-meal correction formula with fat/protein adjustment factor (26); and (e) evidence-based category defaults derived from consensus values in the reference literature.

#### (3) Data-quality correction heuristics

Specific rules for identifying and discarding systematic errors in the reference data, most critically the Diogenes database’s practice of assigning GI = 70 as a placeholder value to non-carbohydrate foods (meats, fats, eggs, condiments, alcoholic beverages). The skill instructs the model to recognize these entries and prefer Sydney International Tables values or nutritional science knowledge instead.

The skill additionally specifies the output format (assigned GI value, match type, reference food, reasoning notes) and an execution procedure. Critically, the skill instructs the model to perform all matching, averaging, and reasoning operations in-context – without writing or executing any programmatic scripts – leveraging the LLM’s capacity for simultaneous semantic understanding and quantitative reasoning across the full reference dataset.

### Validation Study 1: Prediction of the US National GI Database

In the first validation experiment, we assessed the skill’s ability to predict expert-curated GI values for US foods using only European reference data. The skill was configured with the Sydney International Tables and Diogenes database as reference sources, *excluding* the curated US National GI Database (11). The target dataset consisted of 9,428 unique USDA food descriptions from FNDDS versions 1.0 through 2017–2018, as used in our original NHANES analysis. The skill-assigned GI values were compared against the expert-curated final values from Della Corte et al. (11), which served as the ground truth. Expert curation for the US GI Database was performed by KDC, guided by substantive domain expertise in GI methodology and nutritional science (11).

Foods were processed in batches of 250 items per API call, with the full skill definition and complete reference database (∼11,000 entries) loaded into context for each batch. This yielded 38 batches per complete run. Each batch produced approximately 41,000 output tokens and required approximately 12 minutes of processing time, for a total wall-clock time of approximately 7.7 hours per complete run of Validation Study 1.

Performance metrics included: (a) exact match rate (assigned GI = curated GI); (b) close match rate (|assigned GI − curated GI| ≤ 5 GI units); (c) mean absolute error (MAE) across all non-zero-carbohydrate foods; (d) concordance in zero-carbohydrate classification; and (e) distribution of match types and confidence levels. Results were compared with the first-pass performance of the cosine-similarity embedding approach reported in the original publication (75.0% initial AI accuracy, 31.3% retention after manual curation).

### Validation Study 2: Application to European Food Descriptions

In the second validation experiment, the skill was augmented by adding the expert-curated US National GI Database to its reference data (totaling ∼19,000 reference entries). The augmented skill was then applied to 1,157 unique food descriptions representing carbohydrate-containing foods consumed in Mediterranean European populations. Food descriptions consisted of FoodEx2 base terms (simple descriptions) from the EFSA food classification and description system, identified as relevant to dietary glycemic exposure in nationally representative surveys from six Mediterranean countries (27).

Validation was based on direct comparison between the LLM skill-assigned GI values and independent expert GI assignments performed by KDC, a nutritional scientist with expertise in GI methodology. The expert assignments were performed manually and independently of both the skill-based and cosine-similarity approaches, using the same reference literature (Sydney International Tables, Diogenes) and substantive domain expertise. Agreement was assessed using the same metrics as Validation Study 1. Where available, the cosine-similarity embedding algorithm was used as previously described (11) as a secondary comparison benchmark. Foods were processed in batches of 250 items, following the same protocol as Validation Study 1. The European food description set required approximately 5 batches per run.

### Application: Prospective GI Assignment for Recent NHANES Cycles

To demonstrate the practical utility of the validated skill, we applied it prospectively to assign GI values to food codes from recent NHANES cycles not covered by the original US GI Database (11). The USDA periodically updates the FNDDS to reflect changes in the food supply, introducing new food codes and retiring others. We identified all food codes in the most recent available FNDDS versions that were not present in the published 1999–2018 database and applied the augmented skill (Sydney + Diogenes + US GI DB) to assign GI values without manual expert review.

Using the extended GI database, we calculated population-weighted mean dietary GI and energy-adjusted glycemic load (GL) for the newer NHANES cycle(s) using the same analytic approach as Della Corte et al. (11): dietary GI was computed as the carbohydrate-weighted mean of individual food GI values for each participant’s daily intake, and GL was computed as the sum of (GI × available carbohydrate) across all foods consumed. Estimates were weighted using NHANES dietary sampling weights and accounted for the complex survey design. Results were reported alongside the published 1999–2018 trends to assess continuity. For consistency with Della Corte et al. (2024), GI values for compound foods were sourced from the published mixed-meal lookup table (2,550 weighted entries), and per-person dietary GI/GL was computed as the average of NHANES day 1 and day 2 dietary recalls.

### General Procedures

All studies were conducted using Claude Opus 4 (Anthropic, San Francisco, CA), accessed via the Anthropic API, with temperature set to 0 (greedy decoding). Foods were processed in batches of 250 items per API call, with the full skill definition and complete reference database loaded into context for each batch. Each batch produced approximately 41,000 output tokens and required approximately 12 minutes of processing time. Validation Studies 1 and 2 were each run in triplicate under identical conditions to assess reproducibility; concordance metrics (exact agreement rate, near-agreement rate within ±2 GI units, and maximum within-food range) are reported alongside accuracy metrics for each study.

We recorded input and output token counts, wall-clock time, and API cost for each run to enable cost estimation for future applications. All statistical analyses were performed in Python (version 3.11.x). For both validation studies, agreement between the LLM skill and expert assignments was quantified using Pearson correlation, intraclass correlation coefficients (ICC[2,1]; two-way random effects, single measures), mean absolute difference (MAD), and Bland-Altman analysis (bias and 95% limits of agreement). The percentage of assignments within ±5 and ±10 GI units was reported, along with GI category agreement (low [≤55], medium [56–69], high [≥70]) assessed using Cohen’s weighted kappa (κ). Penalized metrics incorporating wrong-zero assignments (expert GI > 0, skill GI = 0) were computed across all expert-positive foods. All tests were two-sided with α = 0.05.

## RESULTS

### Validation Study 1: US National GI Database Prediction

The USDA food database contained 9,428 unique foods with expert-curated GI values, of which 1,455 were assigned GI = 0 (zero-carbohydrate) and 7,972 were assigned GI > 0 by the expert (Table 1).

**TABLE 1.** Validation Study 1 (USDA): accuracy and reproducibility metrics.

| **Metric** | **Value** |
| --- | --- |
| Total foods analyzed (N) | 9428 |
| Expert GI = 0 (n) | 1455 |
| Expert GI > 0 (n) | 7972 |
| **Zero-carb classification** |  |
| Both zero | 1262 |
| Both nonzero | 7711 |
| Expert=0, Skill>0 | 193 |
| Expert>0, Skill=0 (wrong-zero) | 261 |
| Zero/nonzero concordance (%) | 95.2 |
| **Accuracy (both-nonzero foods)** |  |
| n | 7711 |
| Pearson r | 0.68 |
| ICC[2,1] | 0.66 |
| Weighted $\kappa$ | 0.54 |
| MAD (GI units) | 7.5 |
| Mean bias (skill - expert) | -3.2 |
| 95% limits of agreement | -26.5 to +20.2 |
| Within +/-5 GI units (%) | 57.6 |
| Within +/-10 GI units (%) | 73.7 |
| Exact match (%) | 26.8 |
| GI category agreement (%) | 70.9 |
| **Penalized (all expert-positive)** |  |
| n | 7972 |
| MAD including wrong-zeros | 8.8 |
| Within +/-5 including wrong-zeros (%) | 55.7 |
| **Triplicate concordance (all foods)** |  |
| Exact agreement, 3 runs (%) | 53.6 |
| Within +/-2 across runs (%) | 60.9 |
| Within +/-5 across runs (%) | 74.3 |
| Median between-run range | 0 |
| Mean between-run range | 5 |
| 95th percentile range | 25 |

The skill correctly classified 95.2% of foods into the same zero/nonzero carbohydrate category as the expert. The skill assigned GI = 0 to 261 foods (3.3% of expert-positive foods) where the expert had assigned a nonzero value; these were predominantly foods with small but nonzero carbohydrate content at the boundary of the zero-carbohydrate heuristic (e.g., cream-based dips, sauces with small starch contributions). Conversely, the skill assigned nonzero GI values to 193 foods classified as zero-carbohydrate by the expert.

Among the 7,711 foods where both the skill and the expert assigned nonzero GI values, the Pearson correlation was r = 0.68 (ICC[2,1] = 0.66), the mean absolute difference (MAD) was 7.5 GI units, and 57.6% of assignments fell within ±5 GI units of the expert value (73.7% within ±10). The skill exhibited a mean negative bias of −3.2 GI units, with 95% limits of agreement from −26.5 to +20.2 (Figure 1). GI category agreement (low [≤55] / medium [56–69] / high [≥70]) was 70.9% (weighted κ = 0.54). Exact matches (within ±0.5 GI units) were observed for 26.8% of both-nonzero foods. When the 261 wrong-zero assignments were included as penalized errors (all expert-positive foods, N = 7,972), the MAD increased to 8.8.

**FIGURE 1.**
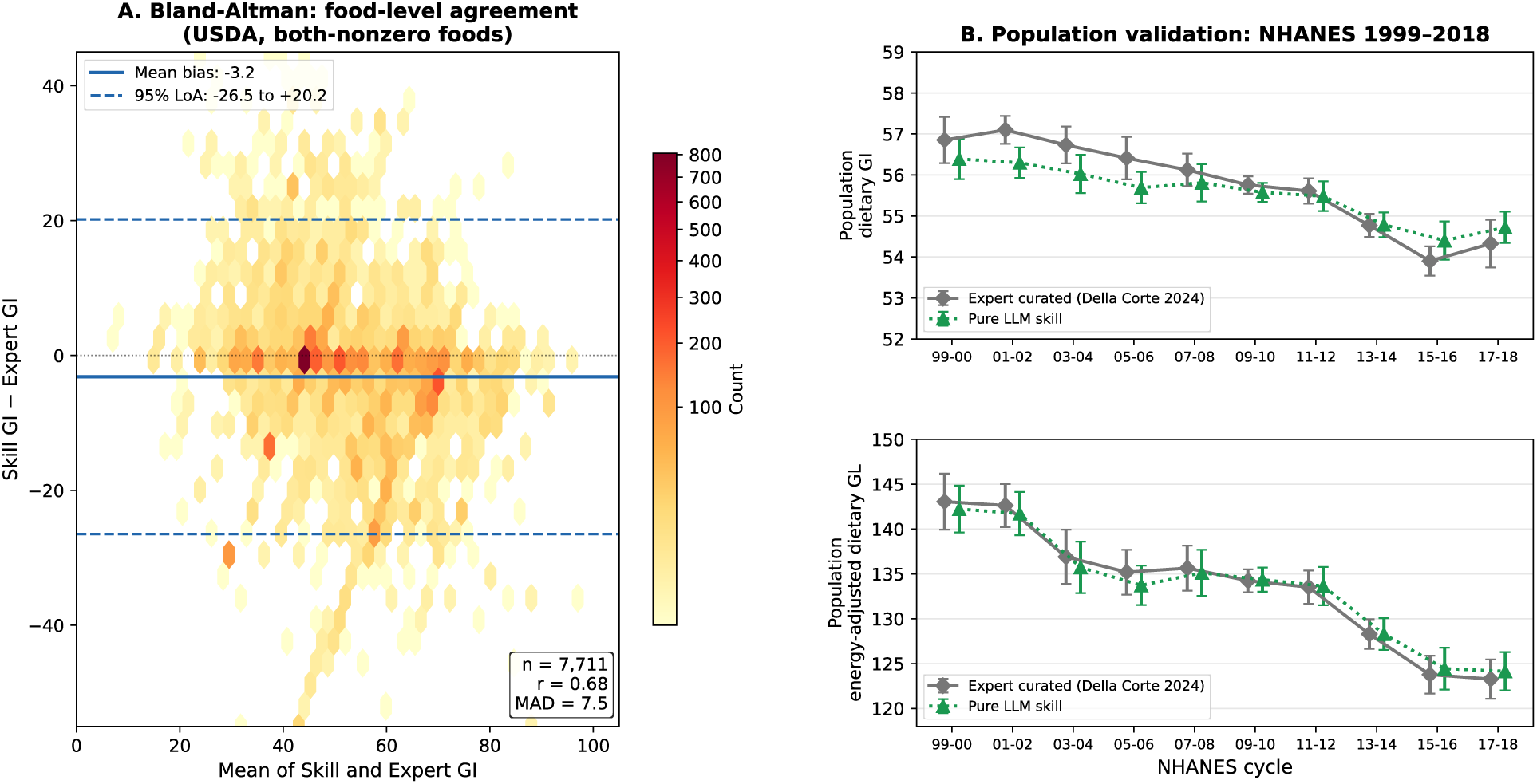
Validation of the LLM skill–based glycemic index (GI) assignment against expert curation, shown at two complementary scales. (A) Bland-Altman density plot of food-level agreement between the skill and the expert-curated US National GI. The hexbin shading (log-compressed via PowerNorm, γ = 1/e) reveals the concentration of foods near zero difference; sparse outliers occupy the periphery. The solid blue line indicates the mean bias (skill − expert = −3.2 GI units); dashed lines indicate the 95% limits of agreement. (B) Population-level validation across the 10 NHANES cycles (1999–2018). Original expert-curated GI assignments (gray diamonds; Della Corte et al. 2024) and skill–assigned reconstruction (green triangles). Error bars represent 95% confidence intervals from the survey-weighted analysis (R survey package; complex sampling design).

These results should be interpreted in the context of two factors. First, this experiment represents a deliberately challenging cross-continental prediction: the skill’s reference data contained only European GI sources (Sydney International Tables and Diogenes), yet the target foods used US-specific food descriptions, preparation methods, and product names not represented in the European reference literature. Second, the expert-curated US GI Database itself required extensive manual refinement – our original cosine-similarity approach achieved 75% initial accuracy but only 31.3% of matches were retained after expert review (11). The skill’s within-±10 agreement of 73.7% on both-nonzero foods, achieved without any US-specific reference data and without manual review, represents a substantial improvement over the first-pass performance of the embedding approach.

To assess whether the food-level scatter observed at the individual food code level (MAD = 7.5 GI units) propagates to population-level dietary surveillance estimates, we performed a parallel reconstruction of the published US dietary GI/GL trends (Della Corte et al. 2024) using exclusively the LLM-skill-assigned GI values, with no expert curation, mixed-meal lookup, or manual review for any food code. Across the 10 published NHANES cycles (1999–2018), the pure LLM skill reproduced the survey-weighted population dietary GI within a mean difference of −0.24 GI units (maximum |Δ| = 0.80) and the energy-adjusted dietary GL within −0.31 units (maximum |Δ| = 1.47) — well within the survey-design 95% confidence intervals (Figure 1B). This demonstrates that the per-food agreement metrics translate to high fidelity at the epidemiologically relevant scale: the LLM, given no US-specific expert input at all, recovers 20 years of national dietary surveillance trends.

Triplicate reproducibility across all 9,428 foods showed 53.6% exact agreement across all three runs, with 60.9% of foods varying by ≤2 GI units and 74.3% by ≤5 GI units between runs. The median between-run range was 0 GI units (mean: 5.0; 95th percentile: 25). A total of 4,365 foods (46.4%) showed some variability across triplicates, reflecting residual non-determinism in LLM inference at scale — even with greedy decoding (temperature = 0), hardware-level floating-point variations and batch context effects can produce different outputs across runs (21).

### Validation Study 2: European Food Descriptions

The European validation set contained 1,157 foods with GI assignments from both the LLM skill and the expert (KDC). Of these, 923 foods received nonzero expert GI values and 234 were classified as zero-carbohydrate by the expert. Metrics including all 1,157 foods are reported in the Supplementary Material (Tables S1–S2).

Among the 907 foods where both the skill and the expert assigned nonzero GI values, the Pearson correlation was r = 0.79 and the intraclass correlation coefficient was ICC[2,1] = 0.79 (Table 2). The mean absolute difference was 6.8 GI units, with 59.2% of assignments within ±5 GI units and 77.2% within ±10 GI units of the expert values. GI category agreement (low/medium/high) was 77.9% (weighted κ = 0.65). These accuracy metrics represent substantial improvements over the cosine-similarity embedding approach evaluated on the same foods (r = 0.65, ICC = 0.64, MAD = 8.8, within ±5 = 53.2%, category agreement = 70.4%, weighted κ = 0.53).

**TABLE 2.**
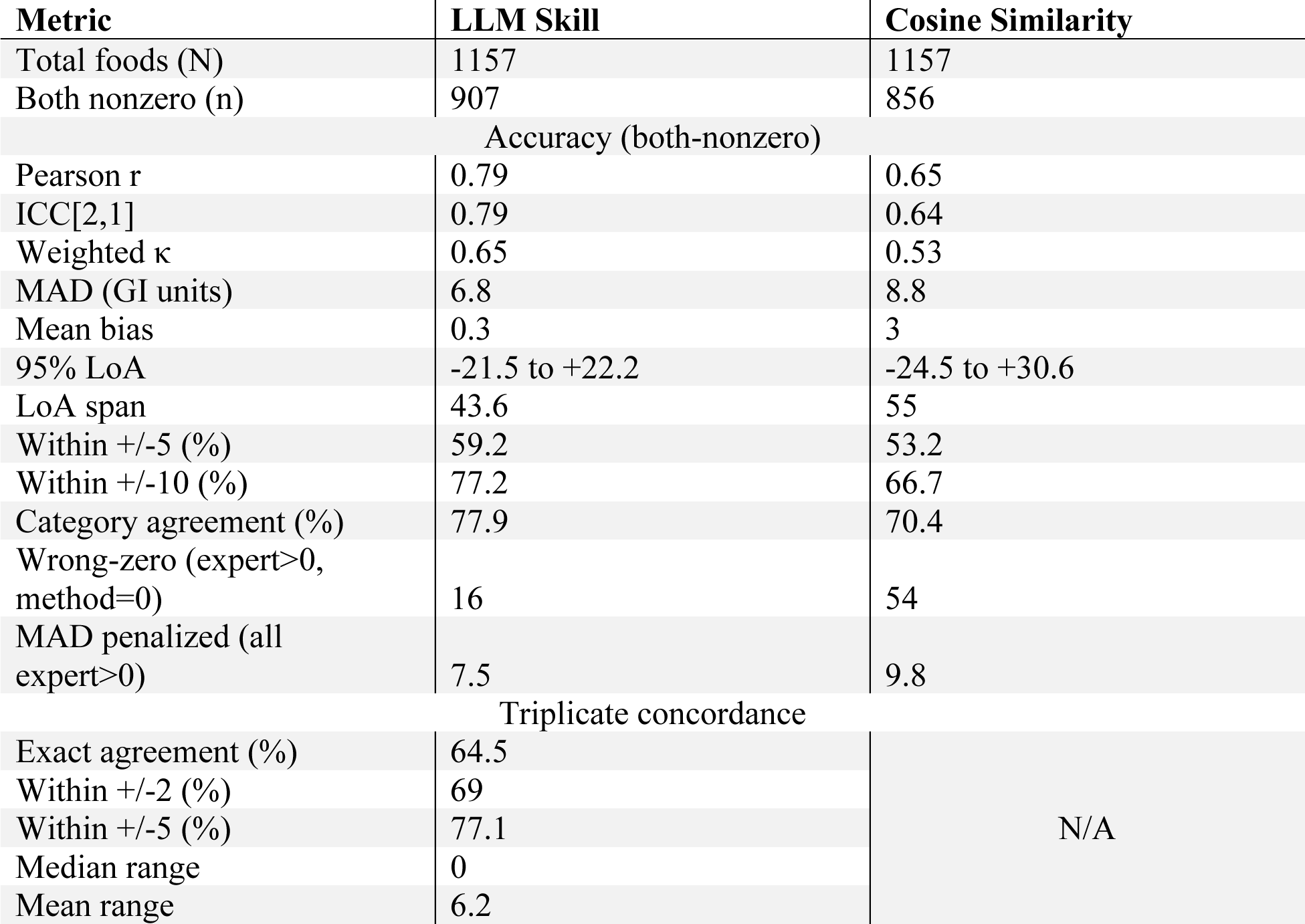
Validation Study 2 (European foods): LLM Skill vs Cosine Similarity.

Bland-Altman analysis revealed a mean bias of +0.3 GI units (skill minus expert) with 95% limits of agreement from −21.5 to +22.2 (span: 43.6 GI units). This was substantially tighter than the cosine-similarity method (bias = +3.0, limits of agreement span: 55.0 GI units) (Figure 2). The skill classified 16 foods (1.7% of the 923 expert-positive foods) as zero-carbohydrate where the expert assigned a nonzero value; when these were included as penalized errors, the MAD increased only modestly to 7.5 (from 6.8), reflecting the low wrong-zero rate.

**FIGURE 2.**
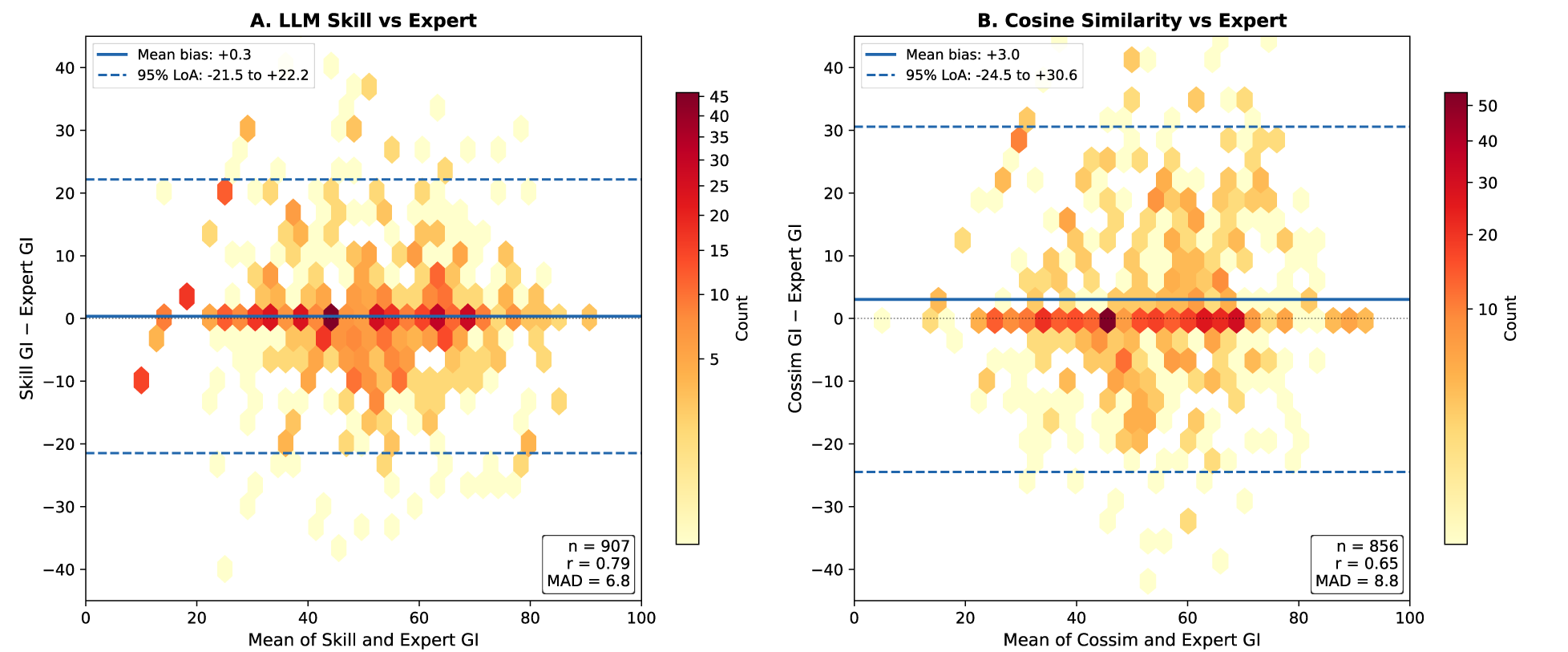
Bland-Altman plots: (A) LLM skill vs expert, (B) cosine similarity vs expert for European foods. LLM skill shows tighter limits of agreement (43.6 vs 55.0 GI units).

Using the triplicate mean of the three skill runs improved agreement slightly (r = 0.79, MAD = 6.8), confirming that averaging reduces noise from stochastic variability. Triplicate reproducibility showed 64.5% exact agreement across all three runs for the full 1,157 foods (triplicate ICC = 0.88), with 69.0% varying by ≤2 GI units and 77.1% by ≤5 GI units between runs (median range: 0; mean: 6.2; 95th percentile: 45).

### Application: Updated US Dietary GI and GL Trends

To extend nationally representative GI and GL surveillance beyond the published 1999–2018 period, we applied the augmented skill (Sydney + Diogenes + US GI DB) to assign GI values to food codes from two additional NHANES cycles: the 2019–March 2020 pre-pandemic cycle (NHANES cycle 11; 12,632 respondents with dietary data) and the August 2021–August 2023 cycle (NHANES cycle 12; 6,751 respondents with dietary data). The latter cycle is the most recently released NHANES data as of this writing. Across both cycles, 410 food codes were identified that were not present in the published US GI Database; the skill assigned GI values to all 410 codes, achieving 100% food code coverage without manual review.

For the 2019–March 2020 pre-pandemic cycle (n = 7,744 adults meeting inclusion criteria), mean dietary GI was 55.0 (95% CI: 54.6, 55.4) and mean energy-adjusted dietary GL was 123.3 (95% CI: 121.3, 125.2). For the August 2021–August 2023 cycle (n = 4,780 adults), mean dietary GI was 55.3 (95% CI: 55.6, 56.0) and mean energy-adjusted dietary GL was 122.0 (95% CI: 120.6, 123.3). These values are reported alongside the published 1999–2018 trends in Figure 3.

**FIGURE 3.**
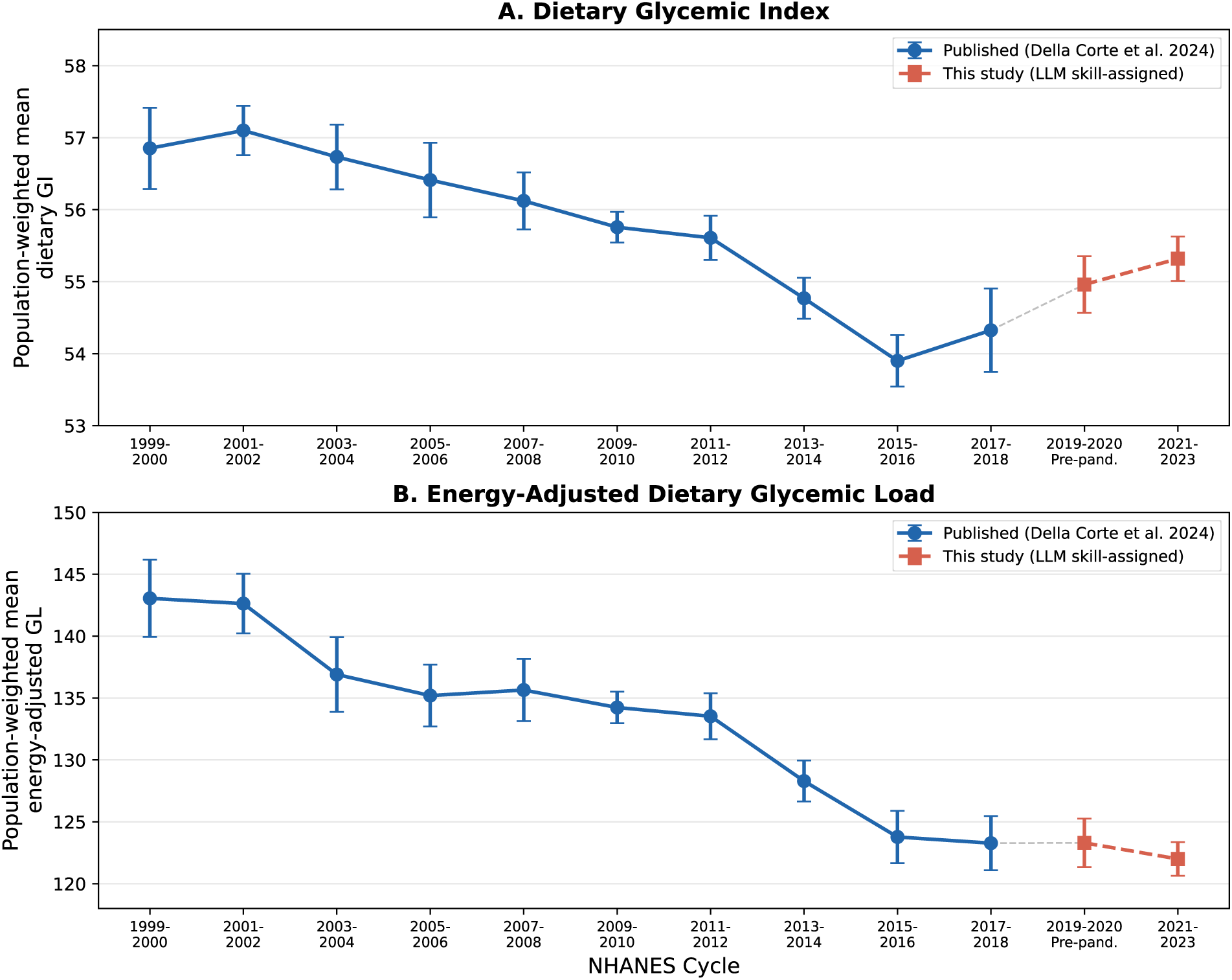
Dietary GI (A) and energy-adjusted GL (B) across 12 NHANES cycles (1999-2023). Blue: published; red: LLM skill-assigned. Survey-weighted using R survey package.

The energy-adjusted GL trend continued its overall decline, decreasing from 143.1 (95% CI: 139.9, 146.2) in 1999–2000 to 122.0 (95% CI: 120.6, 123.3) in 2021–2023, representing a 14.7% decrease over approximately 24 years of surveillance – consistent with the 13.8% decline observed through 2018 in the published data. The dietary GI trend showed a slight uptick from the 2015–2016 nadir (53.9) to the 2019–March 2020 pre-pandemic cycle (55.0) and the 2021–2023 cycle (55.3) but remained well below the 1999–2000 baseline of 56.9. Notably, the August 2021–August 2023 cycle employed telephone-administered dietary recalls (rather than in-person interviews) due to pandemic-related modifications to the NHANES protocol, which may affect comparability with earlier cycles.

The seamless integration of skill-assigned GI values for the 410 new food codes with the 9,428 previously curated values demonstrates the practical utility of the skill-based approach for prospective database maintenance. The entire extension – from identifying new food codes to generating population-weighted trend estimates – required approximately 30 minutes of computation and no expert labor for GI assignment, compared with the months of manual curation required for the original database.

## DISCUSSION

In this study, we developed and validated a skill-based in-context learning approach for assigning glycemic index values to food composition databases. The approach fundamentally differs from previous AI-assisted methods – including our own cosine-similarity embedding algorithm (11) – in that the LLM serves simultaneously as the matching engine, the quantitative aggregation system, and the expert curator, all within a single inference pass. This integration of functions that previously required separate algorithmic and human components is the central methodological advance. More broadly, the skill-based approach can be understood as instantiating a domain-specific knowledge model within the LLM’s context window, enabling integrated reasoning across food composition, GI methodology, and data quality – capabilities that go beyond pattern matching and approach expert-level nutritional reasoning.

### Comparison with Existing Approaches

The skill-based approach addresses the primary limitation identified in our original US GI Database publication (11): the low retention rate (31.3%) of algorithmically generated matches after expert review. Cosine-similarity embeddings excel at identifying textually similar food descriptions but cannot evaluate whether a match is nutritionally appropriate – a determination that requires understanding carbohydrate composition, cooking method effects, and data-quality artifacts. By encoding this expert knowledge directly into the skill, the LLM can reject inappropriate matches (e.g., discarding Diogenes GI = 70 placeholder values for non-carbohydrate foods) and make nuanced substitutions (e.g., selecting an appropriate rice variety based on grain type and preparation method) in a single pass.

Our approach also differs architecturally from retrieval-augmented generation (RAG) systems such as NutriRAG (28), which retrieve a small number of relevant reference entries per query. By loading the entire reference database into context (∼300,000 tokens), the skill enables the model to reason across the full distribution of values for a given food category, compute means across multiple entries, and identify outliers – operations that would require multiple retrieval cycles in a RAG framework. The trade-off is higher per-inference token cost, which we characterize in Table S3.

The FoodSky approach (16) and similar fine-tuned models achieve domain specialization through training on nutrition-specific datasets. The skill-based approach achieves comparable specialization through in-context learning, with two practical advantages: (a) no fine-tuning infrastructure or training data curation is required, and (b) the skill can be instantly updated when new reference data become available (e.g., the next edition of the International Tables) without retraining.

### Reproducibility Considerations

The stochastic nature of LLM inference has been identified as a significant concern for scientific applications (17–21). Our triplicate analysis provides empirical evidence on the magnitude of this concern for structured data-assignment tasks. Our triplicate concordance analysis adopts an aggregate consistency standard rather than exact bitwise reproducibility — analogous to the inter-rater reliability standard used for human expert assessments. Several features of GI assignment constrain the output space in ways that favor reproducibility. First, the task is fundamentally a classification/numerical assignment problem with bounded outputs (GI values 0–100), not open-ended text generation. Second, the reference data are loaded identically in every run, leaving only the model’s matching and reasoning or hardware-level floating-point non-determinism as sources of variability. Third, zero-carbohydrate foods (which typically constitute 40–50% of food codes in national databases) are assigned GI = 0 deterministically based on compositional data, not model reasoning. These constraints substantially reduce the effective stochastic surface area of the task.

To further support reproducibility, the skill architecture is fully described in the Methods section, including the matching hierarchy, category defaults, and data-quality correction heuristics. Combined with a pinned model version and deterministic sampling parameters (temperature = 0), the approach constitutes a well-specified methodological framework. While the complete skill definition is not publicly released – reflecting the substantial expert knowledge and iterative refinement embedded in its construction – the architectural principles and key design decisions are described in sufficient detail to inform the development of analogous skills for other nutritional databases. Researchers requiring GI assignments for new food composition databases are invited to contact the authors for collaboration.

### Practical Implications

The primary practical implication of this work is the dramatic reduction in time and expertise required to construct a national GI database. Our original US database required approximately 6 months of expert labor for manual curation of ∼8,000 food codes (11). The skill-based approach generates a complete set of GI assignments for an equivalent database in approximately 8 hours of computation at a cost of approximately $97 per run ($350 for triplicate validation across all experiments; Table S3). We demonstrated this directly by applying the validated skill to extend the US dietary GI and GL surveillance to recent NHANES cycles without any manual curation, producing trends that are continuous with the published 1999–2018 data. Critically, the food-level scatter observed in Figure 1A averages out almost completely at the population scale (Figure 1B): pure LLM-assigned dietary GI/GL values reproduced 20 years of published NHANES surveillance trends within ∼0.3 absolute units, an error well below the survey-design uncertainty. This suggests that the modest per-food error tolerated by the LLM skill is fully compatible with the precision needed for nutritional epidemiology. This has immediate implications for enabling GI-based dietary surveillance and epidemiologic research in countries and food systems that currently lack GI databases.

The skill architecture is also inherently cumulative: each expert-validated database can be incorporated into the reference data for subsequent applications, progressively improving coverage and accuracy. This is directly demonstrated by the contrast between our two validation studies. In Validation Study 1, the skill achieved ICC = 0.66 and MAD = 7.5 when predicting US foods using only European reference data – a deliberately challenging cross-continental evaluation. In Validation Study 2, after augmenting the reference data with the curated US GI Database, performance on European foods improved substantially to ICC = 0.79 and MAD = 6.8. This progressive improvement mirrors how human expert knowledge accumulates with experience and suggests that the skill will continue to improve as additional validated databases become available.

### Limitations

Several limitations warrant consideration. First, while our reproducibility analysis demonstrates high concordance across triplicate runs, we cannot guarantee identical outputs for every food code across runs – a constraint inherent to all LLM-based methods. We mitigate this by providing concordance rates and recommending triplicate consensus as standard practice. Second, the approach depends on a proprietary LLM accessed via API, introducing dependencies on model availability, version stability, and cost. Future work should evaluate the transferability of this skill to alternative LLMs, including open-source models, to reduce platform dependence. Third, while the skill encodes substantial expert knowledge, it does not replace the need for expert oversight of low-confidence assignments, particularly for novel or highly composite foods. We recommend that skill-based GI assignments be treated as high-quality first drafts subject to targeted expert review, similar to how our original embedding-based approach was used as an initial screen (11). Fourth, both the US GI Database and European food expert assignments were performed by a single expert (KDC); while this expert has demonstrated proficiency in GI methodology through substantive domain expertise (11), future validation with multiple independent raters would further strengthen the evidence.

Fifth, the computational cost of loading ∼300,000 tokens of reference data per inference pass, multiplied across thousands of food codes, is non-trivial. We report exact costs to enable budget planning, and we note that costs are expected to decrease as LLM inference becomes more efficient. Sixth, our validation is limited to US and European food supplies; application to food systems with substantially different food items (e.g., East Asian, African, or South American cuisines) may require augmentation of the reference data with region-specific GI testing. Seventh, the current reference data do not preserve the distinction between Table 1 (foods tested using the standard ISO methodology) and Table 2 (foods tested using non-standard methods) in the International Tables of Glycemic Index Values (6). Expert practice preferentially weights Table 1 values as more reliable; the skill currently treats all reference entries equally regardless of methodological quality tier. Incorporating this quality classification into the reference data is a priority for future versions. Eighth, the skill does not implement region-specific GI selection. Some foods – particularly branded products – have different formulations across markets that produce meaningfully different GI values (e.g., breakfast cereals manufactured with different processing methods in different countries, or soft drinks sweetened with sugar cane vs. high-fructose corn syrup). The current skill averages across all available reference values without regional preference, which can lead to errors for such products.

### Future Directions

Several extensions of this work are warranted. First, application of the skill to national food databases in countries lacking GI data (e.g., developing nations participating in the Global Diet Quality Project) could rapidly expand the geographic scope of GI-based epidemiologic research. Second, the skill architecture is generalizable beyond GI: analogous skills could be developed for assigning other food-composition indices (e.g., NOVA ultra-processing classification, nutrient density scores) that currently require manual expert assignment. Third, integration of skill-based GI assignment into consumer-facing dietary tools – such as meal-planning applications that automatically compute GI values for user-entered meals – could translate this research infrastructure into actionable dietary guidance. Fourth, combining real-time GI assignment with evidence-based substitution thresholds (e.g., recommending lower-GI alternatives when a food exceeds a clinically meaningful GI difference) could enable personalized dietary optimization at scale. Fifth, as LLM context windows continue to expand, future versions of the skill could incorporate additional reference data sources, including continuous glucose monitoring data for personalized GI estimation, manufacturer specifications, and recipe-level ingredient databases.

## CONCLUSIONS

We developed and validated a skill-based in-context learning approach for assigning glycemic index values to food composition databases using large language models. The approach integrates semantic food matching, quantitative reasoning, and expert domain knowledge within a single inference framework, achieving GI assignments comparable to expert ratings across two independent validation databases (US and European). We demonstrated the practical utility of the method by applying it prospectively to extend nationally representative dietary GI and GL surveillance to two additional NHANES cycles (2019–March 2020 pre-pandemic and August 2021–August 2023), assigning GI values to 410 new food codes and producing population-weighted trend estimates that are continuous with the published 1999–2018 data without manual curation. Triplicate concordance analysis demonstrates that the method is reproducible under controlled conditions. The skill architecture and design principles described herein establish a new paradigm for scalable and reproducible GI database construction and maintenance in nutritional epidemiology.

## ETHICS STATEMENT

This study analyzed publicly available, de-identified data from the National Health and Nutrition Examination Survey (NHANES), conducted by the National Center for Health Statistics (NCHS). The NHANES protocol was approved by the NCHS Research Ethics Review Board, and written informed consent was obtained from all participants at the time of original data collection. Because this work involved only secondary analysis of de-identified public-use data, it was exempt from institutional review board review under 45 CFR 46.104(d)(4), and no additional approval was required.

## Data availability

The NHANES dietary data and USDA Food and Nutrient Database for Dietary Studies (FNDDS) food codes analyzed in this study are publicly available from the National Center for Health Statistics (https://www.cdc.gov/nchs/nhanes/) and the USDA Food Surveys Research Group (https://www.ars.usda.gov/northeast-area/beltsville-md-bhnrc/beltsville-human-nutrition-research-center/food-surveys-research-group/docs/fndds/), respectively. Survey-weighted dietary GI and GL estimates for all 12 NHANES cycles (1999–2023), including subgroup-level estimates, are reported in the manuscript and Supplementary Material (Tables S4–S6). Representative food-level results from both validation studies, including triplicate runs, are provided in Tables S1–S2, and example GI assignments for the new NHANES 2019–2023 food codes are listed in Table S7.

The complete skill definition cannot be publicly released because it embeds the full content of the Sydney International Tables of Glycemic Index Values and the Diogenes European food GI database as in-context reference data. These reference sources are the intellectual property of their respective originators and are not available under licenses that permit redistribution. The skill architecture – including the matching hierarchy, zero-carbohydrate rule, data-quality correction heuristics, mixed-meal correction, and category defaults – is described in sufficient detail in the Methods to support independent reimplementation by researchers with access to equivalent reference data. Consistent with our prior work developing the US National GI Database, analysis code and intermediate GI assignment outputs are available from the corresponding author on reasonable request. Researchers wishing to apply the skill-based approach to new food composition databases are invited to contact the corresponding author for collaboration.

## ACKNOWLEDGMENTS

We thank Simin Liu and Carlo La Vecchia for their contributions to the broader Mediterranean GI database project and for reviewing this manuscript.

## AUTHOR CONTRIBUTIONS

The authors’ responsibilities were as follows – **KDC:** conceptualization, data curation (expert GI assignments for both US and European validation databases), validation, writing – review and editing; **JLE**: data aggregation, initial AI prototyping, and manuscript review; **JBM:** manuscript review, validation; **FA:** manuscript review, validation; **DDC:** conceptualization, methodology (skill design and implementation), software, formal analysis, investigation, data curation (reference database preparation), writing – original draft, writing – review and editing, project administration. All authors read and approved the final manuscript.

## SUPPLEMENTARY MATERIAL

The Supplementary Material contains eight supplementary tables and one supplementary figure: Tables S1–S2 present example food-level GI assignment results with triplicate run data for both validation studies (9,428 USDA foods and 1,157 European foods). Table S3 contains the cost analysis.

Tables S4–S5 present survey-weighted dietary GI and energy-adjusted GL by demographic subgroup. Table S6 presents dietary GI and GL across all 12 NHANES cycles (1999–2023). Table S7 lists GI assignments for the 410 new food codes. Table S8 provides worked examples of the skill assignment process illustrating key decision types. Supplementary Material describes the skill assignment workflow.

