## Supplementary Material for "In-Context Learning with Large Language Models for Scalable Glycemic Index Assignment to Food Composition Databases: Development, Validation, and Reproducibility"

Della Corte et al.

### 1. Food-Level Validation Results

#### Table S1. Validation Study 1: USDA Food-Level Results (9,428 foods)

Skill-assigned GI values from triplicate runs versus expert-curated ground truth for 9,428 USDA food codes. The skill used only European reference data (Sydney International Tables and Diogenes). Sample of first 20 foods shown.

| Food Code | Description | Expert GI | Run 1 | Run 2 | Run 3 |
|---|---|---|---|---|---|
| **42101110** | Almonds, salted | 24 | 24 | 24 | 24 |
| **41304970** | Lentils, NFS | 28 | 29 | 26 | 29 |
| **62101100** | Apple, dried | 29 | 29 | 29 | 29 |
| **11830940** | Meal replacement, high protein, milk based, fruit juice mixable | 30 | 30 | 30 | 30 |

| | | | | | |
|---|---|---|---|---|---|
| | formula, powdered, not reconstituted | | | | |
| **41101090** | White beans, NFS | 31 | 31 | 31 | 31 |
| **75204012** | Lima beans, from frozen, no added fat | 33 | 33 | 33 | 33 |
| **91731000** | Peanuts, chocolate covered candy | 33 | 33 | 33 | 33 |
| **11212050** | Milk, evaporated, fat free (skim) | 34 | 34 | 34 | 34 |
| **21103110** | Beef steak, breaded or floured, baked or fried, NS as to fat eaten | 46 | 46 | 46 | 46 |
| **21500200** | Ground beef or patty, breaded, cooked | 46 | 46 | 46 | 46 |
| **22000300** | Pork, NS as to cut, breaded or floured, fried, NS as to fat eaten | 46 | 46 | 46 | 46 |
| **22101400** | Pork chop, battered, fried, NS as to fat eaten | 46 | 46 | 46 | 46 |
| **64105400** | Cranberry juice, 100%, not a blend | 56 | 56 | 56 | 56 |
| **53720200** | Nutrition bar (Clif Bar) | 57 | 57 | 57 | 57 |
| **58123110** | Bao bun | 58 | 58 | 58 | 58 |
| **55300010** | French toast, NFS | 59 | 59 | 59 | 59 |
| **53223000** | Cookie, gingersnaps | 66 | 66 | 66 | 66 |
| **51111010** | Bread, cheese | 67 | 68 | 65 | 68 |
| **53101000** | Cake, angel food, NS as to icing | 67 | 67 | 67 | 67 |
| **55100010** | Pancakes, plain, | 67 | 67 | 67 | 67 |

frozen

#### Table S2. Validation Study 2: European Food-Level Results (1,157 foods)

Skill-assigned GI values from triplicate runs versus expert (KDC) and cosine-similarity algorithm for 1,157 European food descriptions. Sample of first 20 foods shown.

| Food ID | Description | Expert | Cossim | Run 1 | Run 2 | Run 3 |
|---|---|---|---|---|---|---|
| **38097** | Black cherries | 22 | 22 | 22 | 22 | 22 |
| **160489** | Cherries (sweet) | 22 | 22 | 22 | 22 | 22 |
| **160388** | Hazelnuts | 24 | 24 | 24 | 24 | 24 |
| **160375** | Cashew nuts | 25 | 25 | 25 | 25 | 25 |
| **160517** | Plums | 39 | 39 | 39 | 39 | 39 |
| **160365** | Globe artichokes | 45 | 45 | 45 | 45 | 45 |
| **160406** | Milkshakes | 45 | 45 | 45 | 45 | 45 |
| **160413** | Aubergines | 45 | 45 | 45 | 45 | 45 |
| **160419** | Sponge cake | 46 | 46 | 46 | 46 | 46 |
| **37874** | Pizza and similar with vegetable topping | 51 | 70 | 51 | 51 | 51 |
| **160472** | Jam of fruit / vegetables | 51 | 53 | 51 | 51 | 51 |
| **160398** | Common banana | 52 | 52 | 52 | 52 | 52 |
| **38680** | Rice pudding | 59 | 59 | 59 | 59 | 59 |
| **160405** | Sweet potatoes | 61 | 41 | 61 | 61 | 61 |
| **160467** | Kaki | 61 | 63 | 61 | 61 | 61 |
| **160541** | Couscous | 65 | 65 | 65 | 65 | 65 |
| **160601** | Granita | 66 | 66 | 66 | 66 | 66 |
| **160424** | Brioche type products | 70 | 70 | 70 | 70 | 70 |
| **160596** | Flour mix (like wheat/rye/barley/oats and other) | 70 | 70 | 70 | 70 | 70 |
| **160391** | Brown sugar | 71 | 71 | 71 | 71 | 71 |

### 2. Cost analysis

#### Table S3. Computational Cost Analysis

| Experiment | Batches per run | Runs | Output tokens per batch | Time per batch (min) | Cost per run (USD) | Time per run (min) | Total cost (USD) | Total time (min) |
|---|---|---|---|---|---|---|---|---|
| **Validation Study 1** | 38 | 3 | 41411 | 12.1 | 97.26 | 459 | 291.78 | 1378 |

| | | | | | | | | |
|---|---|---|---|---|---|---|---|---|
| **(USDA)** | | | | | | | | |
| **Validation Study 2 (European Foods )** | 5 | 3 | 38275 | 12.1 | 16.82 | 61 | 50.46 | 182 |
| **NHANES Extension** | 2 | 1 | 41094 | 11.1 | 8.25 | 22 | 8.25 | 22 |
| **All experiments** | | | | | | | 350.49 | 1582 |

### 3. Extended NHANES Dietary GI and GL Estimates

#### Table S4. Dietary GI by Demographic Subgroup (1999–2018)

Survey-weighted mean dietary GI by sex, age group, race/ethnicity, education, and income-to-poverty ratio. Computed using the R survey package with 20-year combined weights, accounting for complex survey design (PSU, strata). Inclusion: adults ≥20 y, energy 600–5,000 kcal (males) or 600–4,500 kcal (females), available carbohydrate >5 g.

| Subgroup | N | Mean GI | SE | 95% CI |
|---|---|---|---|---|
| **All** | 49,205 | 55.7 | 0.07 | 55.5, 55.8 |
| **Men** | 23,702 | 56.4 | 0.08 | 56.2, 56.5 |
| **Women** | 25,503 | 55.0 | 0.09 | 54.8, 55.2 |
| **Age20-29** | 8,222 | 56.3 | 0.15 | 56.0, 56.6 |
| **Age30-39** | 8,001 | 55.8 | 0.13 | 55.5, 56.0 |
| **Age40-49** | 7,941 | 55.4 | 0.13 | 55.2, 55.7 |
| **Age50-59** | 7,153 | 55.3 | 0.13 | 55.1, 55.6 |
| **Age60-69** | 7,802 | 55.1 | 0.15 | 54.8, 55.4 |
| **Age>=70** | 8,332 | 55.8 | 0.12 | 55.6, 56.0 |
| **Mexican** | 8,830 | 53.4 | 0.15 | 53.1, 53.7 |
| **Other Hispanic** | 4,003 | 55.4 | 0.18 | 55.1, 55.8 |
| **White** | 21,844 | 55.7 | 0.10 | 55.5, 55.9 |
| **Black** | 10,310 | 57.3 | 0.11 | 57.1, 57.5 |
| **Other Race** | 4,218 | 55.6 | 0.20 | 55.2, 56.0 |
| **Education HS or less** | 23,526 | 56.4 | 0.10 | 56.2, 56.6 |
| **Education Some college** | 13,511 | 55.6 | 0.11 | 55.4, 55.8 |
| **Education College+** | 10,356 | 54.6 | 0.12 | 54.3, 54.8 |
| **PIR <1** | 9,337 | 56.2 | 0.15 | 55.9, 56.5 |
| **PIR 1-2** | 11,974 | 56.2 | 0.12 | 55.9, 56.4 |
| **PIR 2-3** | 6,983 | 56.1 | 0.14 | 55.8, 56.4 |
| **PIR 3-4** | 5,154 | 55.9 | 0.16 | 55.6, 56.2 |
| **PIR 4-5** | 3,684 | 55.2 | 0.19 | 54.8, 55.5 |
| **PIR >=5** | 7,903 | 54.9 | 0.13 | 54.7, 55.2 |

#### Table S5. Energy-Adjusted Dietary GL by Demographic Subgroup (1999–2018)

Energy-adjusted to 2,097 kcal. Same survey design and inclusion criteria as Table S4 above.

| Subgroup | N | Mean dGL | SE | 95% CI |
|---|---|---|---|---|
| **All** | 49,205 | 133.0 | 0.39 | 132.3, 133.8 |
| **Men** | 23,702 | 131.3 | 0.51 | 130.3, 132.3 |
| **Women** | 25,503 | 134.6 | 0.42 | 133.8, 135.5 |
| **Age 20-29** | 8,222 | 138.6 | 0.78 | 137.0, 140.1 |
| **Age 30-39** | 8,001 | 134.5 | 0.79 | 132.9, 136.0 |
| **Age 40-49** | 7,941 | 130.6 | 0.70 | 129.2, 132.0 |
| **Age 50-59** | 7,153 | 128.8 | 0.71 | 127.4, 130.1 |
| **Age 60-69** | 7,802 | 128.4 | 0.68 | 127.1, 129.7 |
| **Age >=70** | 8,332 | 135.1 | 0.47 | 134.2, 136.0 |
| **Mexican** | 8,830 | 129.9 | 0.72 | 128.5, 131.3 |
| **Other Hispanic** | 4,003 | 138.3 | 0.90 | 136.6, 140.1 |
| **White** | 21,844 | 131.6 | 0.54 | 130.5, 132.6 |
| **Black** | 10,310 | 139.4 | 0.65 | 138.2, 140.7 |
| **Other Race** | 4,218 | 137.0 | 0.98 | 135.1, 139.0 |
| **Education HS or less** | 23,526 | 137.7 | 0.48 | 136.8, 138.7 |
| **Education Some college** | 13,511 | 132.0 | 0.52 | 131.0, 133.0 |
| **Education College+** | 10,356 | 126.5 | 0.61 | 125.3, 127.7 |
| **PIR <1** | 9,337 | 140.9 | 0.65 | 139.6, 142.1 |
| **PIR 1-2** | 11,974 | 138.6 | 0.57 | 137.5, 139.7 |
| **PIR 2-3** | 6,983 | 134.6 | 0.73 | 133.2, 136.0 |
| **PIR 3-4** | 5,154 | 132.8 | 0.83 | 131.2, 134.4 |
| **PIR 4-5** | 3,684 | 129.3 | 1.03 | 127.2, 131.3 |
| **PIR >=5** | 7,903 | 124.7 | 0.71 | 123.3, 126.1 |

#### Table S6. Dietary GI and GL by NHANES Cycle (1999–2023)

Per-cycle estimates using 2-year dietary sample weights. Cycles 1–10 (1999–2018) use expert-curated GI values from Della Corte et al. (11). Cycles 11–12 use LLM skill-assigned GI values for 410 new food codes; all other codes retain expert-curated values.

| NHANES Cycle | N | Mean GI (95% CI) | Mean GL (95% CI) | Mean eaGL (95% CI) | GI Source |
|---|---|---|---|---|---|
| **1999–2000** | 4,325 | 56.9 (56.3, 57.4) | 146.0 (141.1, 150.8) | 143.1 (139.9, 146.2) | Published (11) |
| **2001–2002** | 4,828 | 57.1 (56.8, 57.4) | 147.7 (144.0, 151.5) | 142.6 (140.2, 145.0) | Published (11) |
| **2003–2004** | 4,287 | 56.7 (56.3, 57.2) | 140.4 (137.3, | 136.9 (133.9, | Published (11) |

| | | | | | |
|---|---|---|---|---|---|
| | | | 143.5) | 139.9) | |
| **2005–2006** | 4,683 | 56.4 (55.9, 56.9) | 136.4 (132.7, 140.1) | 135.2 (132.7, 137.7) | Published (11) |
| **2007–2008** | 5,459 | 56.1 (55.7, 56.5) | 133.5 (130.3, 136.7) | 135.6 (133.1, 138.2) | Published (11) |
| **2009–2010** | 5,834 | 55.8 (55.5, 56.0) | 133.7 (131.4, 136.1) | 134.2 (133.0, 135.5) | Published (11) |
| **2011–2012** | 4,880 | 55.6 (55.3, 55.9) | 134.6 (132.7, 136.5) | 133.5 (131.7, 135.4) | Published (11) |
| **2013–2014** | 5,086 | 54.8 (54.5, 55.1) | 126.0 (123.7, 128.3) | 128.3 (126.6, 129.9) | Published (11) |
| **2015–2016** | 5,062 | 53.9 (53.5, 54.3) | 120.6 (117.9, 123.3) | 123.8 (121.7, 125.9) | Published (11) |
| **2017–2018** | 4,761 | 54.3 (53.7, 54.9) | 121.1 (118.2, 123.9) | 123.3 (121.1, 125.5) | Published (11) |
| **2019–Mar 2020†** | 7,744 | 55.0 (54.6, 55.4) | 122.7 (120.8, 124.7) | 123.3 (121.3, 125.2) | This study |
| **Aug 2021–2023†** | 4,780 | 55.3 (55.0, 55.6) | 115.8 (114.5, 117.2) | 122.0 (120.6, 123.3) | This study |

†Cycles 11 and 12 use LLM skill-assigned GI values for 410 new food codes. Abbreviations: eaGL, energy-adjusted glycemic load (per 2,097 kcal); CI, confidence interval.

### Table S7. GI Assignments for 410 New NHANES Food Codes

Food codes in the 2019–March 2020 and August 2021–August 2023 NHANES dietary data not present in the published 1999–2018 US GI Database. GI values assigned by the augmented LLM skill without manual expert review. Sample shown.

| Food Code | Food Description | Assigned GI |
|---|---|---|
| **5734400** | Special K | 69 |
| **5816321** | Rice, creamed | 60 |
| **5324800** | Cookie, whole wheat, dried fruit, nut | 55 |
| **5210115** | Biscuit, baking powder or buttermilk type, made fr | 63 |
| **5431300** | Crackers, oyster | 70 |
| **5840900** | Noodle soup, with fish ball, shrimp, and dark gree | 48 |
| **5734410** | Sprinkle Spangles | 70 |

| | | |
|---|---|---|
| **5816331** | Flavored rice mixture | 65 |
| **5816333** | Flavored rice mixture with cheese | 63 |
| **5816335** | Flavored rice, white and wild | 64 |
| **5816336** | Flavored rice, brown and wild | 62 |
| **5816338** | Flavored rice and pasta mixture | 58 |
| **5816341** | Spanish rice | 65 |
| **5816345** | Spanish rice with ground beef | 65 |
| **4120603** | Beans and franks | 48 |
| **5816351** | Rice dressing | 65 |
| **1155105** | Milk fruit drink | 45 |
| **7512100** | Pepper, hot chili, raw | 0 |
| **1155110** | Milk fruit drink, Puerto Rican style (Champola de | 42 |
| **5816361** | Rice-vegetable medley | 65 |
| **7520300** | Bamboo shoots, cooked, fat not added in cooking | 0 |
| **7520302** | Bamboo shoots, cooked, fat added in cooking | 0 |
| **9257010** | Fluid replacement, electrolyte solution | 70 |
| **7430200** | Tomato juice cocktail | 38 |
| **2711610** | Beef curry | 35 |
| **2613313** | Porgy, breaded or battered, baked | 20 |
| **2613314** | Porgy, floured or breaded, fried | 20 |
| **11436100** | Yogurt tube | 33 |
| **2711620** | Beef with barbecue sauce (mixture) | 15 |
| **6111301** | Lemon, raw | 0 |
| **2416711** | Chicken, wing, with or without bone, battered, fri | 20 |
| **2621512** | Turtle (terrapin), cooked, NS as to cooking method | 0 |
| **2736200** | Stewed tripe, Puerto Rican style, with potatoes (M | 48 |
| **5611600** | Noodles, chow mein | 45 |
| **2834513** | Chicken or turkey soup, cream of, prepared with wa | 45 |
| **2834512** | Chicken or turkey soup, cream of, prepared with mi | 45 |

| | | |
|---|---|---|
| **2711635** | Stewed, seasoned, ground beef, Mexican style (Pica | 30 |
| **2834516** | Chicken and mushroom soup, cream of, prepared with | 45 |
| **9257050** | Fluid replacement, 5% glucose in water | 100 |
| **5816411** | Rice with raisins | 65 |
| **5210204** | Biscuit, baking powder or buttermilk type, made fr | 63 |
| **5111901** | Bread, egg, Challah | 70 |
| **5111904** | Bread, egg, Challah, toasted | 70 |
| **9216100** | Cappuccino | 28 |
| **5120101** | Bread, whole wheat, 100% | 69 |
| **5120102** | Bread, whole wheat, 100%, toasted | 69 |
| **5111911** | Bread, lowfat, 98% fat free, toasted | 75 |
| **5111910** | Bread, lowfat, 98% fat free | 75 |
| **5816421** | Rice dessert or salad with fruit | 65 |
| **5120106** | Bread, whole wheat, 100%, made from home recipe or | 65 |

### 4. Skill Assignment Examples

#### Table S8. Examples of Skill-Based GI Assignment Process

Representative examples illustrating how the skill assigns GI values, including cases where the skill corrects errors from the cosine-similarity approach. The skill identifies Diogenes GI=70 placeholder values for non-carbohydrate foods, applies category defaults for unmatched foods, and uses composite reasoning for mixed meals.

| **Food Code** | **Description** | **Expert GI** | **Cossim GI** | **Skill GI** | **Skill Reasoning** |
|---|---|---|---|---|---|
| **14010000** | Cheese, NFS | 0 | 34 | 0 | Zero-carb: cheese has negligible available carbohydrate |
| **21000110** | Beef, NS as to cut, cooked | 0 | 70 | 0 | Zero-carb: pure meat; cossim matched to Diogenes placeholder GI=70 |
| **21601010** | Beef steak, | 0 | 70 | 0 | Zero-carb: |

| | | | | | |
|---|---|---|---|---|---|
| | broiled | | | | pure meat; Diogenes GI=70 correctly rejected |
| **56205010** | Rice, white, cooked | 71 | 63 | 65 | Direct match: white rice in Sydney Tables (mean of multiple entries) |
| **51101010** | White bread, NFS | 72 | 72 | 72 | Direct match: white bread consensus ~72 from International Tables |
| **58102010** | Taco, corn tortilla, beef, cheese | 52 | 52 | 52 | Category match: corn tortilla-based mixed meal |
| **27243010** | Chicken and noodles, cream sauce | 43 | 53 | 48 | Composite: pasta (45) dominant carb, reduced for fat/protein in sauce |
| **57123000** | Oatmeal, regular | 55 | 51 | 55 | Close match: oat porridge from reference (~55) |
| **91746300** | Candy, fruit snacks | NA / New Food Code | 78 | 78 | Category default: candy/gummy confectionery |
| **11300100** | Non-dairy milk, NFS | NA/ New Food Code | 34 | 30 | Category: plant-based milk, similar GI to dairy milk |

Expert GI: final curated value from Della Corte et al. (11). Cossim GI: initial AI-assigned value from cosine-similarity embedding. Skill GI: LLM skill-assigned value (triplicate mean). Empty expert values indicate foods not in the published database.

### 5. Skill Architecture

#### Skill-Based GI Assignment Workflow

The skill-based assignment process operates in a single inference pass per batch of 250 foods. For each food description, the model applies the following decision hierarchy:

1. Zero-carbohydrate check: If the food is a single-ingredient item with negligible available carbohydrate (e.g., unprocessed meat, cheese, oil, water), assign GI = 0. Foods with low but measurable carbohydrate content (e.g., dairy, some fruits, soups) retain their published GI values when available in the reference data.
2. Pre-calculated mixed-meal lookup: If a USDA Food Code match exists in the mixed-meal database (2,550 entries with weighted GI from ingredient-level data), use that value.
3. Direct match: Search the reference database (~11,000 entries) for the same food. If multiple entries exist, compute the trimmed mean excluding outliers.
4. Close match: Match to a closely related food with similar carbohydrate quality, fiber content, and preparation method.
5. Category default: Assign based on food category (28 evidence-based defaults from International Tables consensus values).
6. Composite reasoning: For ambiguous foods, reason about the dominant carbohydrate source and adjust for fat/protein attenuation.

At each step, the skill also applies data-quality correction: Diogenes GI = 70 placeholder values are identified and discarded for non-carbohydrate foods. Energy drinks are assigned GI = 80 (unless sugar-free), glutinous rice products receive GI = 92, and frozen meals are never assigned GI = 0.
